# Chasing the ghost of infection past: identifying thresholds of change during the COVID-19 infection in Spain

**DOI:** 10.1101/2020.04.09.20059345

**Authors:** Luis Santamaría, Joaquín Hortal

**Affiliations:** Estación Biológica de Doñana (EBD-CSIC), C/ Américo Vespucio 26, Isla de la Cartuja, E41092 Sevilla, Spain.; Department of Biogeography and Global Change, Museo Nacional de Ciencias Naturales (MNCN-CSIC). C/José Gutiérrez Abascal 2, 28006 Madrid, Spain.

**Keywords:** Breakpoint regression, Madrid, Catalonia, COVID-19, infection, fatalities, growth curve, social distancing effectiveness

## Abstract

COVID-19 pandemic has spread worldwide rapidly from its first outbreak in China, with different impacts depending on the age and social structure of the populations, and the measures taken by each government. Within Europe, the first countries to be strongly affected have been Italy and Spain. In Spain, infection has expanded in highly populated areas, resulting in one of the largest nationwide bursts so far by early April. We analyze the evolution of the growth curve of the epidemic in both the whole of Spain, Madrid Autonomous Region (the second largest conurbation in Europe), and Catalonia (which includes Spain’s second largest city), based on the cumulative numbers of reported cases and deaths. We conducted segmented, poisson regressions on log-transformed data to identify changes in the slope of these curves and/or sudden shifts in the number of cases (i.e. changes in the intercept) at fitted breaking points, and compared their results with a timeline including both key events of the epidemic and containment measures taken by the national and regional governments. Results were largely consistent in the six curves analyzed (reported infections and deaths for Spain, Madrid and Catalonia, respectively), showing three major clusters of shifts in slopes (growth rates) on March 13-19, March 23-29 and April 1-5 that resulted in 33-71% reductions of slope, and originated in infections on March 3-9, 13-19 and 22-26; as well as a decrease in the infection rate following the strengthened lockdown of 29-30 April, only for Madrid and Catalonia. Small upward shifts in the progress of the disease in Madrid were not associated with significant increases in the intercept of the curve, and seem related with unevenness in case reporting; but they did so in Spain and Catalonia, where they were probably associated to specific events of group infection in Vitoria and to the onset of the outbreak in Catalonia. These results evidence an early deceleration in the spread of COVID-19 coinciding with personal hygiene and social distancing recommendations, as well as the general awareness of the population; and a second, stronger decrease when harder isolation measures were enforced. The combination of these two inflection points seemingly led to the start of the contention of the disease outbreak by early April, the limit of our time series. This highlights the importance of adopting public health strategies that include disseminating basic knowledge on personal hygiene and reduced social contact at the onset of the epidemic, and the importance of early enforcement of hard confinement measures for its subsequent contention.

## Introduction

COVID-19 infection has rapidly spread worldwide since its first outbreak in Wuhan (China) in mid-December 2019. The global number of confirmed cases has gone over one million on 3rd April 2020 (John Hopkins University Coronavirus Resource Center, see Dong et al. 2020), barely 3 months after its first report on 31st December. Individuals infected with COVID-19 remain asymptomatic for 5-6 days, while presenting enough viral load to be infective after 1-2 days of infection (Linton et al. 2020, Lai et al. 2020). Severe cases require hospitalization 3-15 days after the appearance of the first symptoms, which are similar to other infectious respiratory illnesses. This, together with the initial unawareness of the population, led to a high transmission rate of the infection, which spread rapidly to neighboring countries, the Middle East and Europe, and then the rest of the world (see https://nextstrain.org/ncov).

An increasing number of countries was progressively affected, and they responded differently depending on the WHO and local expert advice at the moment, the structure and resources of their public health systems, their R&D capacity (which determined the number of PCRs available for testing contagions from blood samples, among other things), and their ability to implement social distance measures. The diversity of policy responses, together with the preexisting differences in spatial aggregation, social behavior and age structure of their populations, provide a unique array of test cases to understand how different levels and combinations of preventive quarantine and social-distancing measures affected the spread of the pandemic.

COVID-19 arrived to mainland Spain at least in early February (first recorded hospitalization dates back to 15th February; Table S1), if not before (see Deslandes et al. 2020). During February, COVID-19 infection reached Spain repeatedly, mainly via UK and Italy - as evidenced by the presence of fourteen different genetic clusters identified by nextstrain (Hadfield et al. 2018; last accessed 27th April). Different from Italy, where infections were concentrated in the North, the combination of these repeated introductions with early, unnoticed community transmission resulted in consecutive outbreaks in distant, highly populated areas of the Basque Country and Navarra (North), Madrid (Center), Catalonia (North East), Andalusia (South) and Valencia (East) (see timeline in Figure 2 below, and Table S1). This is supported by preliminary evidence for an excess of cases diagnosed as influenza during February and March in Catalonia, compared with the historical record, that may be masking early COVID-19 infections (Coma et al. 2020). The spatial structure of the Spanish populations has played a role in the particularly rapid spread of the pandemic in some regions the country. Its impact has been harsher in the big conurbations of Madrid (around 6.4M people; second most populated Metropolitan area of the EU, after Paris) and Barcelona (c. 5.4M) - as well as in Álava, Navarra and La Rioja (c. 1M in total), following the early infection of healthcare workers from Txagorritxu Hospital. Balearic and Canary archipelagos also received infections from the early onset of the pandemic, so it is reasonable to assume that by early March COVID-19 infections were widely distributed throughout the whole country. The pandemic peaked, however, most strongly in Madrid – to the point that 65-67% of local incidence and mortality at the 52 Spanish provinces is explained by their early-stage mobility from and to Madrid (Mazzoli et al. 2020).

Several factors support the use of Spanish data on the early expansion of COVID-19 to obtain a fair account of the effects of the pandemic at the country and regional levels. Alike Italy and South Korea, early records of the disease (until mid-April, see below) are unbiased, though incomplete. Although the lack of enough tests that has been pervasive for most countries (except South Korea), Spain has achieved one of the highest testing ratios per capita (Clark et al. 2020), thanks to the early mobilization of many PCR machines available in universities and research centers for either COVID-19 testing or COVID-19 research.

Importantly, during this early period only cases testing positive in the PCR make it to the official statistics, and (similar to Italy but different to other European countries) all deaths testing positive were registered as caused by COVID-19 infection, including those associated with previous pathologies. Until mid-April, these data excluded many deaths happening outside hospitals (e.g. in private homes and nursing homes), which were not tested. From this date onwards the nearly twofold increase in the testing capacity, as well as the inclusion of previous cases and deaths diagnosed from symptoms but not tested, created a serious unevenness in the time series. Although this update of cases is providing an increasingly more realistic account of the extent of the epidemic, it creates a temporal bias in reporting that hampers the fair comparison of data before and after mid-April. Therefore, we restrict our analyses to data from February, March and the first two weeks of April 2020. These data provide an underestimation of the total population infected and the number of fatalities, due to the limited number of tests (although for the number of deaths, this is partly compensated by the lethality associated to other pathologies that is attributed to COVID-19 when PCRs render positive tests). However, the relatively homogeneous intensity of testing and the stability of criteria for disease attribution throughout the time period of this analysis probably result in unbiased estimators for the spread of the pandemic. It is therefore safe to assume that the number of reported cases of infection and the number of deaths during this early period are reasonably good proxies for the advance of the pandemic.

Here we characterize the growth curve of COVID-19 infections in the whole of Spain, from the onset of the pandemic in early February through the establishment of increasingly more restrictive social and governmental restrictions to mobility and personal contact. We also perform the analyses for the Madrid and Catalonia Autonomous Regions (Madrid and Catalonia hereafter), which represent the country’s two largest foci of the pandemic. Madrid is a highly populated area with good public transportation and a high daily commuting rate, while Catalonia combines Barcelona metropolitan area (the second largest in the country) with an extensive rural territory of low population density. Therefore, they represent prime examples of the spread of the virus in large sets of populations through time, mostly panmictic in the case of Madrid and spatially heterogeneous in the case of Catalonia, as well as of the effect of social-distancing measures thereupon.

The adoption of containment measures by the national and regional governments followed a sustained increment through time, from (1) the recommendation of preventive measures in late February and early March, to (2) increasingly stricter social-distancing measures on 9-10 March, to (3) a nationwide lockdown announced on 13 March and enforced on 15 March, to (4) an strengthened lockdown, with the closure of all non-essential economic activities on 31 March (see Figure 2 below, and Table S1). Non-essential economic activities were allowed again from 13th April, coinciding with the last value of the time series analyzed here. Since infections are not detected as cases until several days later (see below), our data contains only a progressive strengthening of contention measures, without any de-escalation. Such sequence of measures was broadly discussed by experts, media and social networks, with opinions ranging from qualifying them as exaggerated or unnecessary, during the first weeks of the outbreak; to criticizing them as tardy of insufficient, in the weeks that followed. Two controversies have been particularly strong: (i) were preventive and soft social-distancing measures useful, or should hard social-distancing measures have been introduced from the early moments (late February to early March)?, and (ii) did the mass events on the weekend of 7-8 March, coinciding with the International Women’s Day demonstrations (over 300k attendants in the whole country, 120K in Madrid and 50K in Barcelona) and premier football league matches (around 280k spectators in total and 72K and 77K in Madrid and Barcelona, respectively) trigger the early spread of the pandemic in Spain’s largest cities, especially in Madrid?

Bearing this temporal sequence in mind, we analyze the growth curves of the cumulative numbers of cases detected and the cumulative number of deaths for the whole of Spain and both Madrid and Catalonia, focusing specifically in the changes in the growth rate (i.e. the slope of log-transformed data) of these curves through time. Based on this analysis, we seek to answer two questions: (1) how effective were the different social-distancing measures in reducing infection and mortality rates?; and (2) how significant were the effects of 7-8 March mass gatherings on the expansion of the epidemic, compared with other key events and control measures?

## Data and methods

### Timeline of events and control measures

Data on the different events that marked the evolution of the pandemic in Spain (e.g. first cases detected, large infection bouts, first deaths) or influenced its perception by the general public, as well as policy measures (e.g. preventive isolation, social-distancing, lockdowns) and putative key events (e.g. large gatherings associated to sport events, political demonstrations and party rallies), were gathered from official sources, national and international media, and scientific publications. Whenever possible, and in all cases for policy measures, we confirmed their date and content from official documents and/or websites from international, national or regional institutions. We include a broad list of events in Supplementary Table S1 and selected the most relevant ones for the timeline shown, together with the results of the statistical analyses, in Figure 2’s graphical summary.

### Infection and fatality data

Official data on the (i) cumulative number of cases, and (ii) cumulative number of deaths were obtained from the daily Covid reports of the Spanish Ministry of Health. Data were extracted at two levels of aggregation, for Spain as a whole country, and for Madrid and Catalonia Autonomous Regions (i.e. *Comunidades Autónomas de Madrid* and *de Catalunya)*. For the analyses we included data from the first day in which at least 10 cases or at least 1 death were measured; and extended the analyses to 29-31 days after the onset of social-distancing measures on 13-15/3/20, a period tripling the average infection-to-detection time (10.1 days; see next section), and significantly larger than the average infection-to-death time (21 days; see next section). This ensures that the effects of these first measures are fully included in the dataset, and also that, in the case of reported cases, data covers the effects of the most stringent confinement measures starting the 31st of March.

### Lag time estimates

To estimate the infection date of reported cases, we calculated the infection-to-testing time by combining reported values of incubation time (mean = 5.0 days in Lauer et al. 2020; median = 5.1 days in Linton et al. 2020; mean = 6.4 days in Lai et al. 2020) with time from illness onset to hospital admission for treatment and/or isolation (median = 3.3 days among living cases and 6.5 days among deceased; Linton et al. 2020). Hence, we used an infection-to-testing time of 9 days for living cases and 12 days for dead cases. Based on the proportion of 36% deaths to 64% recoveries reported from 3/3/20 to 6/4/20 (for a total of 57,006 closed cases in Spain), we estimated an average infection-to-testing time of 10.1 days – which, for simplicity, was rounded to 10 days. Similarly, to estimate infection date from day of death, we combined the reported values of incubation time (mean = 5.0 days in Lauer et al. 2020; median = 5.1 days in Linton et al. 2020; mean = 6.4 days in Lai et al. 2020) and time from illness onset to death (mean = 15.0 days in Lauer et al. 2020; mean = 15 days in Linton et al. 2020, as used also by Russell et al. 2020; mean = 17.8 in Verity et al. 2020) - which resulted in a infection-to-death time of 21 days.

### Analyses

We fitted a family of segmented (broken-line) regressions with no, one, two, three, four and up to five breaking points (Models 0 to 5, with two, four, six, eight, ten and twelve parameters respectively) and compared them using their respective AICs, using the segmented and lme4 packages of R 3.6.3 (R Core Team 2020). We chose the segmented package because, different to R’s strucchange package (Zeileis et al. 2002) and other ‘structural breaks models’, it requires the fitted lines to join at the estimated breakpoints (i.e. it results in nearly-continuous models), which is consistent with the type of data analyzed. However, we also conducted additional analyses using strucchange, to increase the probability of identifying additional breakpoints (see below). Models with a Relative Likelihood >0.05 were considered as equally good to that with the lowest AIC; we report primarily on the most parsimonious of these models (i.e., the model with less breaking points), but include also a brief discussion of all comparable models. Since we are analyzing count data, the model with zero breaking points (and all segmented models built upon it) was fitted using Generalized Linear Models, with a poisson error distribution and a log link.

For each model, we used the residual deviance and its degrees of freedom to calculate the scale (thus identifying residual overdispersion, if present) and evaluate the model’s fit. While both values showed a marked improvement as the model fit improved, they did not reach scale values significantly similar to 1 in all cases. Given the limitations of a strict use of the p-values for the evaluation of model goodness-of-fit in poisson regression (e.g. Bartlett 2014), the strong improvement in both scale and test values in the better models (i.e. those with lower AICs), and the fact that increasing further the number of breaking points resulted in model instability, we are however confident that our models are as good as this technique allows.

Fitted breaking points provide objective information on the moment at which infection dynamics changed, while slopes provide information of the direction and magnitude of such changes. When analyzing the data from Madrid, we observed discontinuities that suggested that some breakpoints could involve a change in the intercept, rather than in the slope. This would imply a significant shift in values at a given day, followed by a continuous increase at the same growth rate that preceded such day – an scenario consistent, for example, with a sudden increase in infection rate during the mass gatherings of 7-8 March. To test for this possibility, and to use a second approach that was more sensitive to events in the early part of the curve, we fitted a second set of segmented regressions using the breakpoints function of R’s strucchange package (Zeileis et al. 2001) with a minimum of five data points per segment. While segmented uses an algorithm that minimizes the increment in the fitted function at breakpoints, thus approaching continuity, strucchange’s breakpoints uses separate intercepts at each different segment, thus allowing for discontinuities in the fitted functions - such as the “jumps” (and, potentially, “stalls” due e.g. to reporting delays) described above. This comes at a cost in terms of increased number of parameters: one more per breakpoint (hence, models 1 to 5 have 3, 6, 9, 12 and 15 parameters, respectively), making the procedure more conservative in terms of breakpoint detection. Moreover, because this procedure can only be applied on ls models and we had to use ln-transformed y-values to ensure linearity, the residuals showed considerable heteroscedasticity - with larger variance for larger x-values. This means that the fitting procedure was more sensitive to variation in the lower part of the x (time) range, i.e. it tended to overestimate the number of breakpoints at the beginning of the curve and underestimate them at the end - as compared to the segmented fits. Altogether, the combination of both fitting procedures provides a more robust testing of the hypothesis presented above: breakpoints fitted in both procedures are likely to be the most relevant for the data series, and those detected by one of the procedure are complementary in terms of the methods’ sensitivity.

## Results

### Number of cases

For the whole of Spain, the model with five breaking points (Model 5) provided the best fit (Table 1). Fitted breaking points were placed on day 18.5 (13-14/3/20, estimated infection on 3-4/3/20), 23.9 (19/3/20, estimated infection on 9/3/20), 31.5 (26-27/3/20, estimated infection on 16-17/3/20), 36.8 (1/4/20, estimated infection on 22/3/20) and 41.3 (5/4/20, estimated infection on 26/03/20) (Figure 1). The growth rate of the number of cases decreased by 42% (from 0.36 to 0.21) after the first breakpoint (13-14/3/20), by 19% (to 0.17) after the second (19/3/20), by 44% (to 0.09) after the third (26-27/3/20), by 38% (to 0.06) after the fourth (1/4/20) and by another 46% (to 0.03) after the fifth (5/4/20).

**Table 1.**
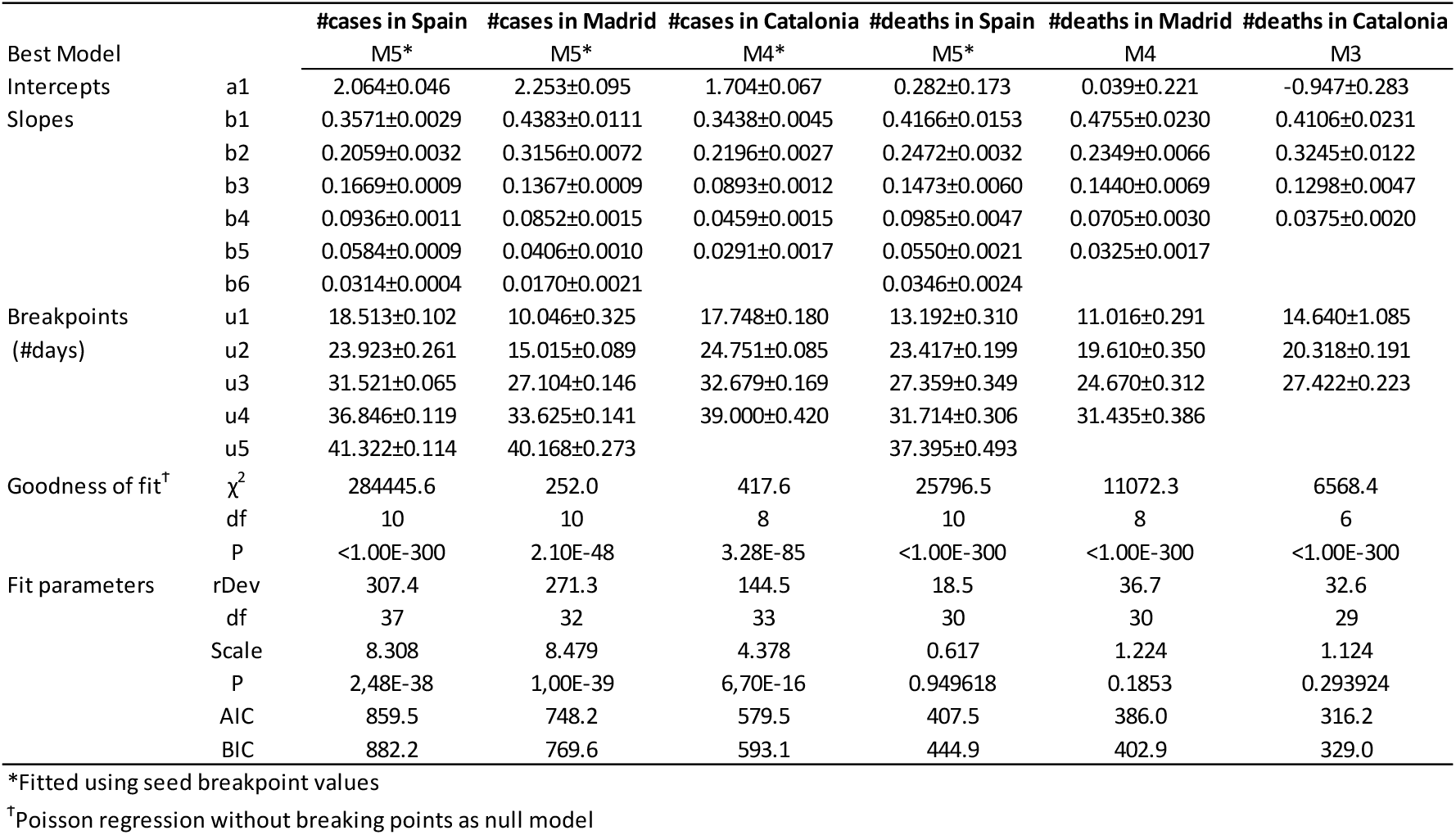
Results of segmented regressions with increasing numbers of breaking points, fitted on the total number of cases and the total number of deaths reported in Spain and the two autonomous regions hosting its two largest cities (Madrid and Catalonia) from 25/2/20 to 13/04/20. Within this period, data series varies among variables and regions, since they start at the first day with >10 cases or >1 death. rDev: residual deviance.

**Figure 1.**
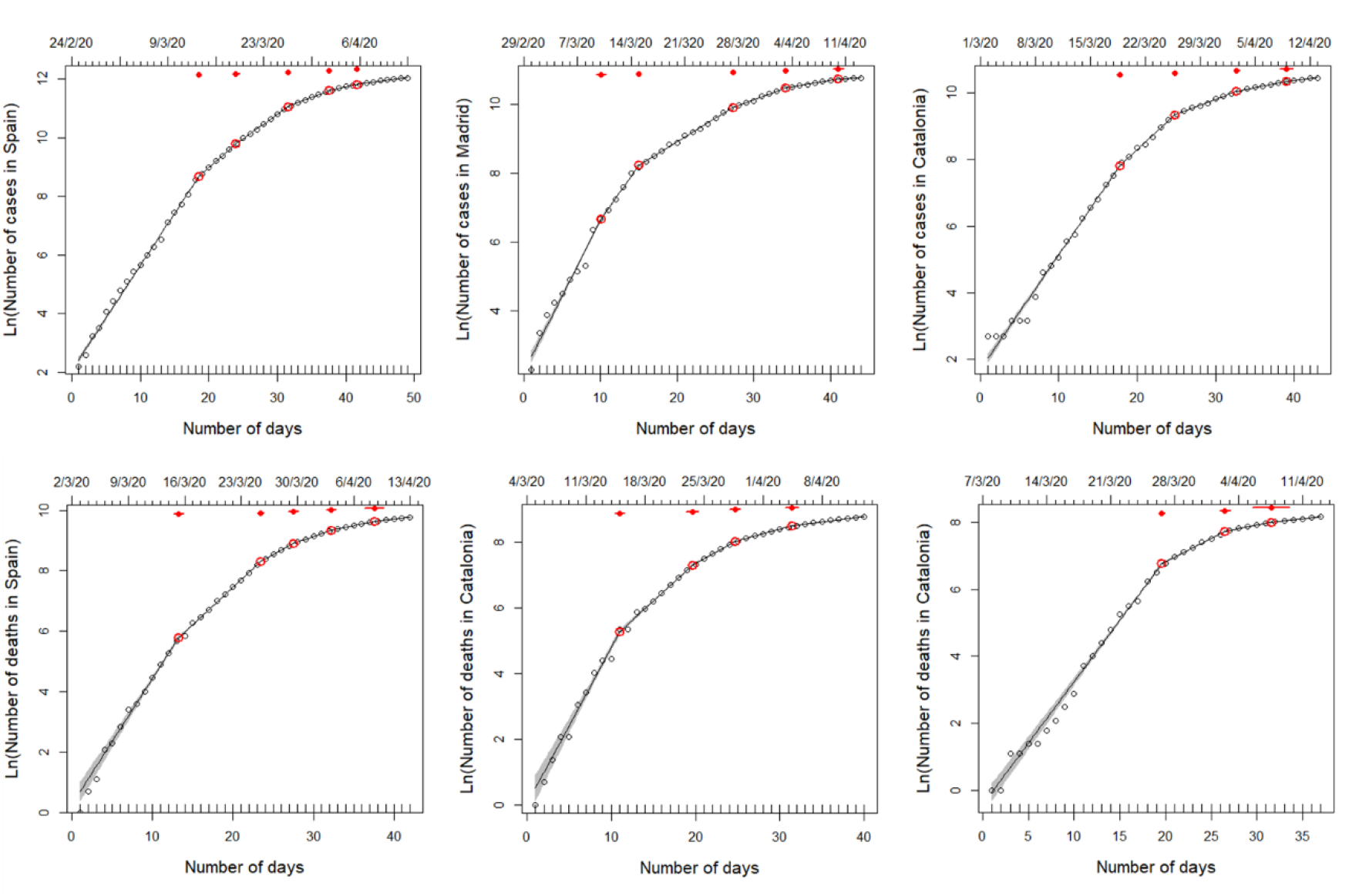
Segmented regressions fitted on the total number of cases detected in Spain and the two autonomous regions hosting its two largest cities (Madrid and Catalonia) from 25/2/20 to 13/04/20. Within this period, data series varies among variables and regions, since they start at the first day with >10 cases or >1 death (see upper X-axis for initial date). Lines show the best fit, as specified in Table 1 (see also Tables S2-S3). Red dots indicate breaking points of the best fit, with 95% confidence intervals (red lines).

The analyses performed on the number of cases from Madrid are consistent with the results for the whole country. The model with five breaking points (Model 5) provided the best fit (Table 1). Fitted breaking points were placed on day 10.0 (10/3/20, estimated infection on 29/2/20), 15.0 (15/3/20, estimated infection on 5/3/20), 27.1 (27/3/20, estimated infection on 17/3/20), 33.6 (3/4/20, estimated infection on 24/3/20) and 40.2 (9/4/20, estimated infection on 30/3/20) (Figure 1). The growth rate of the number of cases decreased by 28% (from 0.44 to 0.32) after the first breakpoint (10/3/20), by 57% (to 0.14) after the second (15/3/20), by 38% (to 0.08) after the third (27/3/20), by 52% (to 0.04) after the fourth (3/4/20) and by another 58% (to 0.02) after the fifth (9/4/20).

Results for Catalonia are largely consistent with those from the whole country and Madrid, although they show a lower number of breaking points. The model with four breaking points (Model 4) provided the best fit (Table 1). Fitted breaking points were placed on day 17.7 (19/3/20, estimated infection on 9/3/20), 24.7 (26/3/20, estimated infection on 19/3/20), 32.7 (3/4/20, estimated infection on 24/3/20), and 39.0 (9/4/20, estimated infection on 30/3/20) (Figure 1). The growth rate of the number of cases decreased by 36% (from 0.34 to 0.22) after the first breakpoint (19/3/20), by 59% (to 0.09) after the second (29/3/20), by 49% (to 0.05) after the third (3/4/20), and by another 37% (to 0.03) after the fourth (11/4/20).

In general, the results of the alternative set of segmented regressions fitted with the strucchange package support those described above. Both procedures consistently identified the 2-3 main breakpoints of each curve (i.e. March 12-15, 19-20 and 25-29, corresponding to estimated infections on March 2-5, 9-10 and 15-19) and showed comparable decreases in the growth rates at all of them (47-67%, 37-48% and 60-67% respectively). As expected, strucchange failed to detect the two breaking points fitted by segmented at the end of the curve (on April 1-5 and 8-11), but it rather identified a significant increase in slope coupled to an increase in the intercepts (i.e., a ‘jump’ on the curve) on the early stages of the outbreak at Spain and Catalonia (on March 6-9, estimated infection time on February 25-28), but not in Madrid. This increase in growth rate was fairly small (19%) in Spain (where it was preceded by a larger decrease, of 37%, one week earlier), but very large in Catalonia (118%), where the start of the curve had been nearly flat until these dates. In Madrid, the apparent stall-and-jump of the number of cases visible on the two first weekends of March (28/2-1/3 and 7-9/3/20) did not result in any significant breakpoint in the strucchange fit.

### Number of fatalities

The model with five breaking points (Model 5) provided the best fit (Table 1) for the fatalities associated to COVID-19 infection in the whole of Spain. Fitted breaking points were placed on day 13.2 (15/3/20, estimated infection time on 24/2/20), 23.4 (25/3/20, estimated infection on 4/3/20), 27.3 (29/3/20, estimated infection on 8/3/20), 31.7 (3/4/20, estimated infection on 13/3/20) and 37.4 (8/4/20, estimated infection on 18/03/20) (Figure 1). The growth rate of the number of fatalities decreased by 41% (from 0.42 to 0.25) after the first breaking point (15/3/20), decreased by 40% (to 0.15) after the second (25/3/20), decreased by 33% (to 0.10) after the third (29/3/20), decreased by 44% (to 0.05) after the fourth (3/4/20), and decreased again by another 37% (to 0.03) after the fifth (8/4/20).

The analyses on the number of fatalities from Madrid are consistent with the nationwide results. The model with four breaking points (Model 4) provided the best fit (Table 1). Fitted breaking points were placed on day 11.0 (15/3/20, estimated infection on 24/2/20), 19.6 (24/3/20, estimated infection on 3/3/20), 24.7 (29/3/20, estimated infection on 8/3/20), and 31.4 (4/4/20, estimated infection on 14/3/20) (Figure 1). The growth rate of the number of cases decreased by 51% (from 0.47 to 0.23) after the first breakpoint (15/3/20), decreased by 39% (to 0.14) after the second (24/3/20), decreased by 51% (to 0.07) after the third (29/3/20), and decreased again by 54% (to 0.03) after the fourth (4/4/20).

In the case of Catalonia, however, the curve of accumulated fatalities showed a lower number of segments, than in Madrid and all Spain. The model with three breaking points (Model 3) provided the best fit (Table 1). Fitted breaking points were placed on day 14.6 (22/3/20, estimated infection on 1/3/20), 20.3 (27/3/20, estimated infection on 6/3/20) and 27.4 (3/4/20, estimated infection on 11/3/20) (Figure 6). The growth rate of the number of cases increased by 21% (from 0.41 to 0.32) after the first breakpoint (22/3/20), decreased by 60% (to 0.13) after the second (27/3/20), and decreased again by 71% (to 0.04) after the third (3/4/20).

The results of the alternative set of segmented regressions fitted with the strucchange package supported generally those described above. Both procedures consistently identified the 2-3 main breakpoints of each curve (i.e. March 12-15 and 25-29, and April 2-4, corresponding to estimated infections on February 20-23, March 3-6 and 12-14) and showed comparable albeit lower decreases in the growth rates at all of them (41-43%, 39-60% and 44-72% respectively). As expected, strucchange identified one additional breakpoint at the beginning of each of the three curves (on March 7-12, estimated infection time on February 15-20), while failing to detect approximately half of them (four out of nine) at the end of the three curves. The two early breaking points detected on March 12-13 and 17-18 in Catalonia corresponded to increases in the intercepts (a ‘dive’ and a ‘jump’ in values, both from one day to the next), with virtually no change in the slopes. In Madrid, two similar jumps, detected on March 6-7 and 12-13 (estimated infections on February 15-16 and 20-21), were associated in contrast with considerable decreases (32-43%) in the growth rate.

## Discussion

The use of segmented regressions provided an objective procedure to identify thresholds of change during the evolution of COVID-19 pandemic in the whole of Spain and the regions containing its two largest cities, Madrid and Catalonia. The results of the six variables analyzed, involving three different spatial settings (whole country; Madrid’s large, panmictic conurbation; and Catalonia urban-rural, heterogeneous region), and two different lag times (10 days for cases, 21 days for fatalities), showed a consistent temporal pattern (Figure 2) that can be divided into five consecutive phases:

1. A first phase, in the early moments of the epidemic in Spain, characterized by fast and sometimes sudden increases in the number of infections (i.e. ‘jumps’). These jumps are particularly conspicuous in Catalonia, and in general for the number of fatalities. This phase coincided with the detection of the first cases, imported from abroad; and the two jumps in the number of infections were synchronous with specific events of group infections (at sport events, first, and hospitals, nursing homes and churches, later; red points 4, 6 and 8, yellow points 1-2, blue point 2 in Figure 2; see also Supplementary Table S1 and Supplementary Figures S1). Owing to the lag times mentioned above, the effects of this phase become perceivable between 1st and 8th March. During this phase, however, a breaking point signaling a 41-51% decrease in the growth rate of the number of deaths was identified at both Spain and Madrid (observed on 15/3/20, estimated infection on 23/2/20, Fig.1). This breaking point is probably related to the improvement of clinical procedures (detection and hospital treatment) following the issuing of specific guidelines for the identification, treatment and research of Covid19 cases by the Spanish Ministry of Health on 19-20/2/20.
2. A second phase, 1-2 weeks later, showing a highly consistent pattern of decreases in the growth rates of both the number of cases and the number of fatalities at the three geographical extents (Spain, Madrid and Catalonia) corresponding to infections starting on March 3-6 (49-57% and 39-60% reductions for cases and deaths, respectively). This cluster of breakpoints is the second most consistent and supported, coinciding closely in time for five out of six of the growth series evaluated, and present in all models with two or more breakpoints, except for the number of cases in Madrid, and in the alternative fits using strucchange segmentation method. This suggests that this decrease was one of the two most important changes of dynamic during the period of time analyzed, despite not being associated with any active confinement measures. Rather, the diminution in the rate of infections in the first week of March seemingly follows the issuing of preventive isolation measures by the Labor Ministry, guidelines for Covid treatment at hospitals and ICUs by the Health Ministry, and other measures by central and regional agencies (red points 7 and 9 in Figure 2) and the increased awareness of the public opinion (as the perception of the increases in cases and deaths described in phase 1 emerged). Importantly, it clearly precedes the issuing of most social-distancing measures by the central and regional governments (‘NL’ in Figure 2). Owing to the lag times mentioned above, these decreases become perceivable between March 14-15 (for the number of cases) and March 24-29 (for the number of deaths). Strikingly, the perception of strong slowdown in March 14-15, broadly understood in the Spanish media as a reflection of the success of the social distancing measures, was actually caused by the combined impact of two sets of previous events: the reduction in the number of infections that started in March 4-5, and the reduction in the number of deaths that started during phase 1 (for infections taking place after February 23) - underscoring the decoupling of cause and perception (see below).
3. A third phase, starting only a few days later, showing a more inconsistent set of reductions for cases (for Spain and Catalonia) and deaths (for Spain and Madrid). This cluster of breaking points corresponds to infections on March 8-9, and resulted in smaller reductions for cases and deaths (19-36% and 33-51%, respectively). The most likely cause is the issuing of partial social-distancing measures by the central and regional governments (purple point 4 and yellow point4 in Figure 2).
4. A fourth phase, starting 5-10 days later, showing a consistent pattern of decrease in the growth rates of the number of deaths (44-71% decrease, estimated infections on 13-14/3/20) and the number of cases (38-59% decrease, estimated infections on 17-19/3/20) at the three spatial extents (Spain, Madrid and Catalonia). This cluster of breakpoints is both the most consistent and the most supported, coinciding for all six growth curves and present in all models with two or more breakpoints, in and most models with one, and in the alternative fits using strucchange. This indicates that this was the most important change of infection dynamics during the period of time analyzed. It coincides closely with the issuing of strong social-distancing measures (nationwide lockdown and border closure on 13-14/3/20), albeit showing a small (3-5 days) delay for the number of cases. The impact of this sharp change on infection trend was only visible on May 27-29 and April 3-4 (for the number of cases and deaths, respectively). This phase includes a third cluster of breakpoints in the number of cases, caused by a reduction in the number of infections starting on April 22-26, which does not seem directly related to any specific event or policy measure - and is most likely related to the strengthening of the lockdown as it was being progressively enforced at local levels.
5. In contrast with other measures - which resulted with highly consistent clusters of breaking points at the three spatial levels of analysis (albeit more asynchronous in Catalonia), the strengthened lockdown issued on March 30th only reduced the number of cases in Madrid and Catalonia, but had no perceivable effect in the whole of Spain. Given the synchrony in time and its limited extent, this reduction might be related primarily to the emergency actions taken in the nursing homes - which concentrated 50-70% of the mortality in these two regions. Unfortunately, the data series is not prolonged enough to allow for an evaluation of the effect of the strengthened lockdown on the number of deaths.

**Figure 2.**
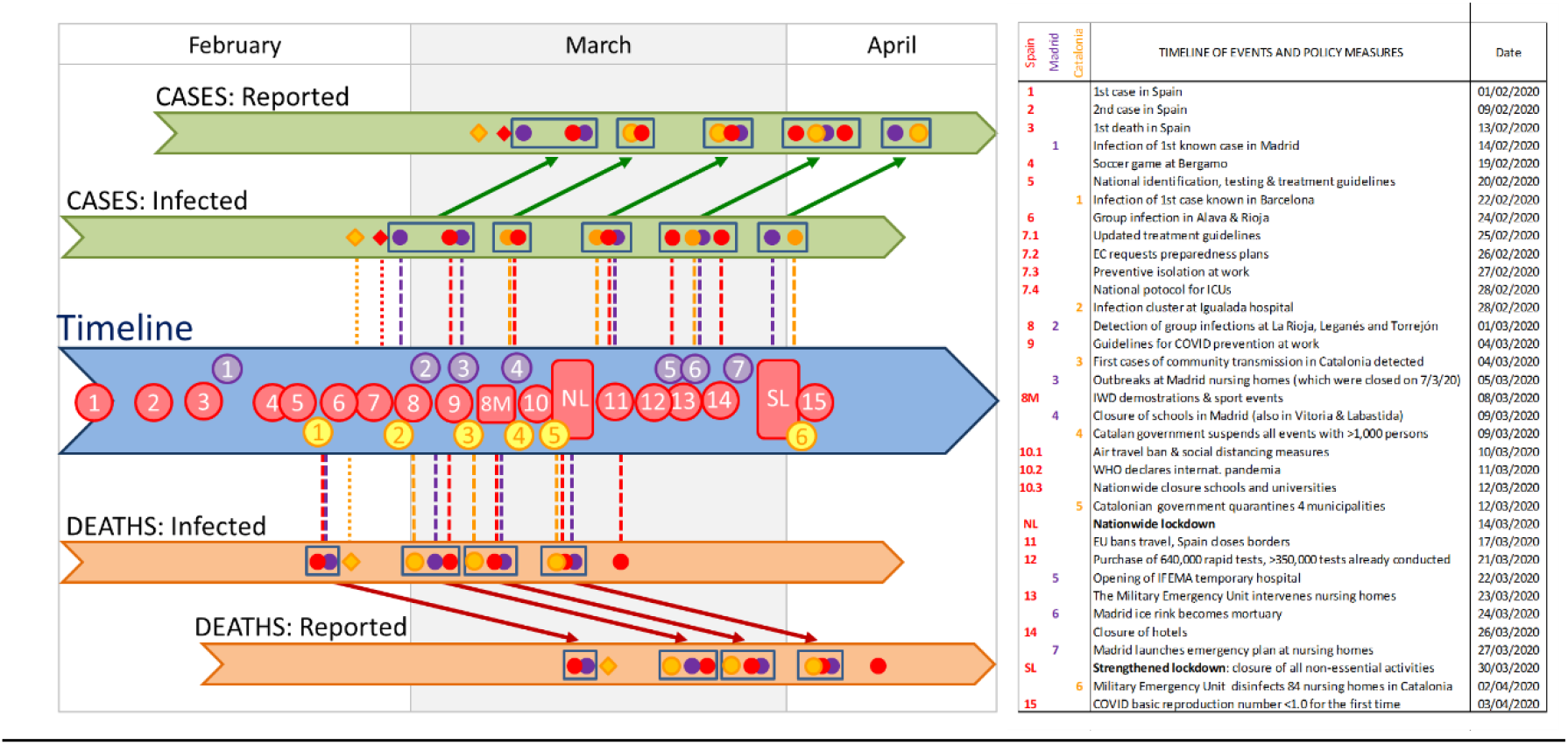
Timeline of the key events for the spread of COVID-19 in Spain, the increased awareness of the Spanish population, and Control Measures taken by the government. Colored dots indicate the breaking points identified in our segmented analyses for the whole of Spain (red) and specifically for Madrid (purple) and Catalonia (yellow). Diamonds indicate sudden increases in the intercept, identified as breaking points by the strucchange analysis. Clusters of breaking points are identified with rectangles. The position of these breaking points in the reported sections indicates the date detected in our analyses of the temporal COVID-19 growth curves, and those in the infected sections indicate the estimated date of infection. Numbers in the timeline indicate the key events listed in the table. Vertical lines indicate potential synchronies between events in the timeline and estimated changes in the growth rates (i.e., breaking points) of both cases and deaths (above and below the timeline, respectively). Diagonal arrows link the estimated infection dates and the detection dates of identified breaking points, for both cases and deaths. Note that the combination of vertical and diagonal lines indicates the dates at which changes in infection dynamics could be perceived, for the first time, as breaking points in the numbers of cases or deaths. WMC stands for World Mobile Congress, and EC for European Commission. See Supplementary Table S1 for a detailed account of the timeline, and Supplementary Figure S1 for an account including the variation in the growth rate (slope of the ln-transformed data) at each breaking point.

This basic analysis of the growth rates of cases and fatalities evidences the effectiveness of the contention measures taken by Spanish national and regional governments. The confinement of all population that could telework, the suspension of in-class teaching in schools and universities, and the closure of non-essential shops, bars and restaurants resulted in a marked downwards inflection in the curves of both infections and deaths. However, it is also remarkable the existence of an earlier breakpoint, corresponding to a decrease in the rate of infections in the first week of March. The dropdown in the rate of infections coincides with an increase of the awareness of the Spanish population (due to the reporting of a rapidly increasing number of cases and deaths, i.e. the delayed perception of the events of phase 1) and the issuing of official recommendations for the prevention and treatment of COVID infections, but precedes the legal enforcement of most social-distancing measures by the regional and central governments (see Figure 2, Table S1). Indeed, it takes place the week before the marked decrease in community mobility registered during the second week of March (from March 8 to March 15) in Madrid, Catalonia and the whole of Spain (Google 2020). This suggests that a large proportion of the Spanish population changed their behaviour due to the combination of increasing awareness and the dissemination of preventive hygienic and distancing measures by the government and the media – with recourse to the examples of China, South Korea and Italy. It seems therefore fair to argue that, during this week, the population responded swiftly to the voluntary prevention and distancing measures suggested by the authorities and the media, and these were effective to slow down the early spread of the pandemic. Indeed, it provides evidence for the potential effectiveness of controlling the initial phases of growth of this pandemic by establishing ‘soft’ contention measures based on public health policies and enhanced use of the primary health care system.

The results also indicate, however, that these measures were not enough to contain the spread of the virus – and the issuing of the first soft social-distancing measures by the regional governments had only a moderate impact upon it (phase 3). The next breaking point, resulting in the strongest reduction in the growth rate of the number of cases, took place closely after the onset of the stronger measures of national lockdown and border closure. As in the previous phase, the population’s response was swift and started already with the partial confinement measures, one week before (e.g. Google 2020) - greatly surpassing the expectations of 69 experts who predicted a collapse of the health system on March 25th (Catanzaro 2020; Arenas et al. 2020; Mitjà et al. 2020). These stronger isolation policies were probably instrumental in flattening the curve of infections, though not sufficient to bring it to a complete halt.

What our results fail to show are the expected effects of any of the key events pinpointed by the media on the spread of COVID-19 infections. The demonstrations and football matches of 7-8 March did not result in significant increases in neither infection rates nor in the number of infections (as reflected in the numbers of cases and fatalities). An inspection of the stalling-and-jump of the numbers of cases and deaths in Madrid, which triggered such public perception, indicates that they are inconsistent with it – since, given the lag times involved, their effects should have shown up ten to 21 days later, not immediately after it. Moreover, the only breakpoints that could be consistent with a change in the number of infections in March 8 (those on March 14-15 for the number of cases in Spain and Catalonia, and March 28-29 for the number of deaths in Spain and Madrid) actually show a decrease in the infection rates - instead of the expected increase in either the slope or the intercept. Instead, the stalling-and-jump pattern, which can also be observed in the previous and following weekends (1st and 15th March, for the number of cases and the number of deaths, respectively), is more likely related to the phase 1 infection bouts shown in Figure 2 and Table S1 (such as Valencia’s soccer game in Bergamo and the group infections of Alava’s funeral, Torrejón’s and Leganes’ churches, and Valdemoro’s nursing home). Alternatively, they could be caused by droppings in patient attendance to hospitals or case/fatality reporting during the weekend, with the subsequent increase in the following Monday.

With hindsight, it is clear that issuing strong social-distancing measures earlier would have increased their effectiveness, thereby saving more lives and reducing the collapse of the Spanish health system (see e.g. the SIR-based simulations of Casares & Kahn 2020). Our analyses evidence, however, that the responses of the population, media and authorities were slowed down by the perceptual trap created by COVID-19’s prolonged infection-to-detection and infection-to-death lags. It is somehow ironic that the rapid increase in the infections during the last weeks of February (described in phase 1) was only perceivable two weeks later - and, while the increments in number of cases and deaths were attributed to concurrent events that were most likely unrelated, they triggered a swift response precisely at a moment when early containment measures were already starting to work. Similarly, the results of early containment measures were perceived two weeks later, and attributed to a direct China, South Korea and Italy. It seems therefore fair to argue that, during this week, the population responded swiftly to the voluntary prevention and distancing measures suggested by the authorities and the media, and these were effective to slow down the early spread of the pandemic. Indeed, it provides evidence for the potential effectiveness of controlling the initial phases of growth of this pandemic by establishing ‘soft’ contention measures based on public health policies and enhanced use of the primary health care system.

The results also indicate, however, that these measures were not enough to contain the spread of the virus – and the issuing of the first soft social-distancing measures by the regional governments had only a moderate impact upon it (phase 3). The next breaking point, resulting in the strongest reduction in the growth rate of the number of cases, took place closely after the onset of the stronger measures of national lockdown and border closure. As in the previous phase, the population’s response was swift and started already with the partial confinement measures, one week before (e.g. Google 2020) - greatly surpassing the expectations of 69 experts who predicted a collapse of the health system on March 25th (Catanzaro 2020; Arenas et al. 2020; Mitjà et al. 2020). These stronger isolation policies were probably instrumental in flattening the curve of infections, though not sufficient to bring it to a complete halt.

What our results fail to show are the expected effects of any of the key events pinpointed by the media on the spread of COVID-19 infections. The demonstrations and football matches of 7-8 March did not result in significant increases in neither infection rates nor in the number of infections (as reflected in the numbers of cases and fatalities). An inspection of the stalling-and-jump of the numbers of cases and deaths in Madrid, which triggered such public perception, indicates that they are inconsistent with it – since, given the lag times involved, their effects should have shown up ten to 21 days later, not immediately after it. Moreover, the only breakpoints that could be consistent with a change in the number of infections in March 8 (those on March 14-15 for the number of cases in Spain and Catalonia, and March 28-29 for the number of deaths in Spain and Madrid) actually show a decrease in the infection rates - instead of the expected increase in either the slope or the intercept. Instead, the stalling-and-jump pattern, which can also be observed in the previous and following weekends (1st and 15th March, for the number of cases and the number of deaths, respectively), is more likely related to the phase 1 infection bouts shown in Figure 2 and Table S1 (such as Valencia’s soccer game in Bergamo and the group infections of Alava’s funeral, Torrejón’s and Leganes’ churches, and Valdemoro’s nursing home). Alternatively, they could be caused by droppings in patient attendance to hospitals or case/fatality reporting during the weekend, with the subsequent increase in the following Monday.

With hindsight, it is clear that issuing strong social-distancing measures earlier would have increased their effectiveness, thereby saving more lives and reducing the collapse of the Spanish health system (see e.g. the SIR-based simulations of Casares & Kahn 2020). Our analyses evidence, however, that the responses of the population, media and authorities were slowed down by the perceptual trap created by COVID-19’s prolonged infection-to-detection and infection-to-death lags. It is somehow ironic that the rapid increase in the infections during the last weeks of February (described in phase 1) was only perceivable two weeks later - and, while the increments in number of cases and deaths were attributed to concurrent events that were most likely unrelated, they triggered a swift response precisely at a moment when early containment measures were already starting to work. Similarly, the results of early containment measures were perceived two weeks later, and attributed to a direct consequence of the national lockdown. Fortunately, expert advice to the government was undoubtedly aware of this perceptual trap and insisted on the necessity of stronger social-distancing measures –which have been narrowly sufficient, to date, to reach the objective of flattening the curves.

The spread and growth intensity of the COVID-19 pandemic is being driven by international connectivity (Coelho et al. 2020), which resulted in repeated exposition of the highly interconnected European population. In the case of Spain, this has been particularly important at international hubs such as Madrid or Barcelona, although contagions due to tourism and sport events also initiated the spread at least in Malaga and Valencia (southern and eastern coasts, respectively), importing the disease at multiple sites and facilitating community transmission throughout the whole Spanish territory (see Supplementary Table S1). This implies that the risk of new introductions from abroad will regain importance, should the objective of flattening the curve and taming the outbreak succeed in Spain. Our results, however, underscore the importance of the prolonged delays between infection and detection for the early (re)containment of the infection. Though population awareness and widespread adoption of preventive measures will likely slow down the advance of the disease in the future, this does not diminish the importance of implementing a much more active and complete system of early detection. This represents a formidable challenge for Spain, whose public health and R&D systems (the latter being instrumental for supporting testing efforts) were severely hit by the post-2008 austerity measures.

The analyses described above are highly consistent, and they were not strongly affected by changes in neither the extension of the data series (as more data became available) nor the fitting procedures (glm vs ls regressions). However, their validity is potentially constrained by a number of caveats. We used data on the number of reported cases – i.e. severe cases subject to testing. In general, tests were restricted to those requiring hospitalization or belonging to risk groups, thus underestimating total infection numbers. Similarly, while at the beginning the number of deaths recorded in Spain included all deaths of patients who tested positive or showed a compatible symptomatology, during the peak of the pandemic most regional governments failed to count all deaths outside hospitals if they were not tested either pre- or post-mortem. Given the significant numbers of deaths happening in nurse homes and particular residences, this unevenness in the measurement of COVID-related fatalities could have biased results, eventually flattening the curve. If such effect was significant, however, it should have resulted in changes in the intercept of the number of deaths’ curve. Given the lack of evidence for such changes, we believe that our results will stand out if new data coming from autopsies or re-evaluations is available in the future. Indeed, we stop our analyses of the time series when these cases started to be reported at an uneven rate by the regional governments, by mid-April, precisely to avoid spurious errors in the detection of breaking points. These limitations emphasize that, at present, we can only use the available data as proxies for the actual rates of infection spread and lethality. Therefore, our results must be taken with due caution until more detailed studies surveying seroprevalence in the general populations are available (including the broad study launched by the Spanish government as we write; Spanish Ministry of Health, 2020).

It is also important to note that the relationship of the growth rates and breakpoints in the analyzed data with the timeline provided in Figure 2 may be affected by two factors: the variability in the time lags between the time of infection and the times of both diagnosis and death (see introduction), and the irregular reporting of both variables during weekends and among Spanish regions –particularly at the onset of the outbreak. However, the use of cumulative curves smoothers out considerably these effects, and provides robust estimates of both breaking points and growth rates. Finally, the use of a family of models reaching a maximum of five breaking points, necessary to maintain model parsimony, may have also resulted in the lack of detection of additional breaking points. However, all models showed highly significant fits (as compared with the reference model, without breaking points); and for half of the studied time series (three out of six curves), the best model had less than five breaking points. We might have also missed breaking points in the early stages of the infection, when values are lower and data noisier. These breaking points were indeed more readily detected by our alternative fitting method (using R’s strucchange package), but should be taken into account with caution given the risk of overfitting related to the higher number of parameters per breakpoint used by such method.

To summarize, our analyses detected five clusters of breakpoints-including two major inflection points-in the dynamics of the COVID-19 pandemic in Spain: a first phase of rapid infection, accompanied by a decrease in the mortality rate most likely related to early improvements in the detection and treatment of this novel disease; a second phase of early reduction in the infection rate, chronologically related to public response to voluntary prevention and social-distancing requests by the authorities, followed by a second response to compulsory measures of partial social-distancing; and a third phase of major reduction in the infection rate, following the onset of hard social-distancing measures (nationwide lockdown and land border closure) and its subsequent strengthening. It is also apparent that prolonged infection-to-detection and infection-to-death lags may have caused misperceptions of both the effectiveness of certain measures and the impact of certain events (most notably, the 8th March demonstrations and several sport events), while being probably instrumental in triggering the widespread social response during phase 2 of the outbreak in Spain. Therefore, in addition to the control measures undertaken, a more effective communication strategy that bridges the perceptual gap described, in combination with socially- and geographically-specific actions carried out by enhanced public health and primary health care systems, could be instrumental in engaging the population into the harsh actions still necessary to fight the pandemic more efficiently.

## Data Availability

All data used in the article are publicly available in public sourced provided in the the Methods section. They are also provided in Supplementary Table S4.

https://www.worldometers.info/coronavirus/country/spain/

## Acknowledgements

Critical comments by Miguel A. Rodríguez-Gironés, David Alonso and Joaquín Calatayud on an early version of the manuscript are gratefully acknowledged.

